# Severe COVID-19 is associated with fungal colonization of the nasopharynx and potent induction of IL-17 responses in the nasal epithelium

**DOI:** 10.1101/2022.10.25.22281528

**Authors:** Carly G. K. Ziegler, Anna H. Owings, Vincent N. Miao, Andrew W. Navia, Ying Tang, Joshua D. Bromley, Peter Lotfy, Meredith Sloan, Hannah Laird, Haley B. Williams, Micayla George, Riley S. Drake, Yilianys Pride, George E. Abraham, Michal Senitko, Tanya O. Robinson, Michail S. Lionakis, Alex K. Shalek, Jose Ordovas-Montanes, Bruce H. Horwitz, Sarah C. Glover

**Author notes:** Correspondence to: Jose Ordovas-Montanes, Bruce H. Horwitz, Sarah C. Glover, and Alex K. Shalek. these authors contributed equally (Carly G. K. Ziegler, Anna H. Owings). these senior authors contributed equally (Alex K. Shalek, Jose Ordovas-Montanes, Bruce H. Horwitz, Sarah C. Glover).

## Abstract

Recent case reports and epidemiological data suggest fungal infections represent an under-appreciated complication among people with severe COVID-19. However, the frequency of fungal colonization in patients with COVID-19 and associations with specific immune responses in the airways remain incompletely defined. We previously generated a single-cell RNA-sequencing (scRNA-seq) dataset characterizing the upper respiratory microenvironment during COVID-19, and mapped the relationship between disease severity and the local behavior of nasal epithelial cells and infiltrating immune cells. Our study, in agreement with findings from related human cohorts, demonstrated that a profound deficiency in host immunity, particularly in type I and type III interferon signaling in the upper respiratory tract, is associated with rapid progression to severe disease and worse clinical outcomes. We have now performed further analysis of this cohort and identified a subset of participants with severe COVID-19 and concurrent detection of *Candida* species-derived transcripts within samples collected from the nasopharynx and trachea. Here, we present the clinical characteristics of these individuals, including confirmatory diagnostic testing demonstrating elevated serum (1, 3)-β-D-glucan and/or confirmed fungal culture of the predicted pathogen. Using matched single-cell transcriptomic profiles of these individuals’ respiratory mucosa, we identify epithelial immune signatures suggestive of IL-17 stimulation and anti-fungal immunity. Further, we observe significant expression of anti-fungal inflammatory cascades in the nasal and tracheal epithelium of all participants who went on to develop severe COVID-19, even among participants without detectable genetic material from fungal pathogens. Together, our data suggests that IL-17 stimulation – in part driven by *Candida* colonization – and blunted type I/III interferon signaling represents a common feature of severe COVID-19 infection.

## INTRODUCTION

Infection with SARS-CoV-2, the virus that causes COVID-19, can lead to severe viral pneumonitis and the development of acute respiratory distress syndrome.^1,2^ Severe COVID-19 is characterized by peripheral immune dysregulation, and we and others, have previously demonstrated blunted interferon responses within the nasal mucosa of patients with severe COVID-19.^3–5^ Recent case reports and retrospective cohort studies suggest secondary infection with fungal pathogens may be a significant contributor to morbidity and mortality in patients with severe COVID-19.^6–11^ The frequency of fungal colonization of the airways in patients with severe COVID-19 and potential impact on local mucosal immunity remains unknown.^9,12–15^ IL-17, released by CD4 T cells and innate lymphocytes, is a key effector cytokine that coordinates mucosal anti-fungal immunity among other adaptive and innate leukocytes, granulocytes, and mucosal stroma.^16–19^ Recent work has uncovered complex interactions between IL17-driven inflammation, type 1 interferon responses, and susceptibility to fungal pathogens, however the effect of fungal colonization and anti-fungal immune responses during co-occurrent SARS-CoV-2 infection have yet to be explored^20^. Here, using a previously-published dataset derived from a cohort of individuals acutely infected with SARS-CoV-2, we directly assessed co-occurrent fungal colonization in the airways of patients with severe COVID-19 and examine pathways associated with anti-fungal immunity.^3^

## RESULTS

We had previously described a cohort of 58 individuals – 56 of which are further characterized here – including 15 healthy participants, 35 individuals diagnosed with acute COVID-19, and 6 intubated patients that were negative for SARS-CoV-2.^3^ Nasopharyngeal (NP) swabs obtained from these patients were employed in a cross-sectional study of the nasal respiratory cellular composition using single-cell RNA sequencing (scRNA-seq) (**Figure 1A, 1B**). Patients in this cohort with COVID-19 were sampled within 9 days of hospital admission (median: hospital day 2), which we estimated was within 2 weeks of initial respiratory symptoms. Full cohort demographic data and findings relating to the cellular composition, behaviors, and response to SARS-CoV-2 infection between disease groups can be found in our prior manuscript. We had previously applied a meta-transcriptomic taxonomic classification analysis to each sample to assign both cell-associated and ambient/extracellular sequencing reads to a reference database of human and microbial genomes (generated on 5/5/2020 from the NCBI Reference Sequence Database including archaeal, bacterial, viral, protozoan, and fungal genomes).^21,22^ This approach, in addition to direct reference-based alignment, enabled us to quantify respiratory abundances of SARS-CoV-2, and connect viral abundances to the cellular sources of viral replication, as well as concurrent epithelial host responses **(Figure 1B)**. Here, we describe further analysis of the data generated from these nasopharyngeal samples, as well as additional data generated from matched endotracheal aspirates (ETA) obtained from 4 of the individuals in the original cohort with severe COVID-19.

**Figure 1.**
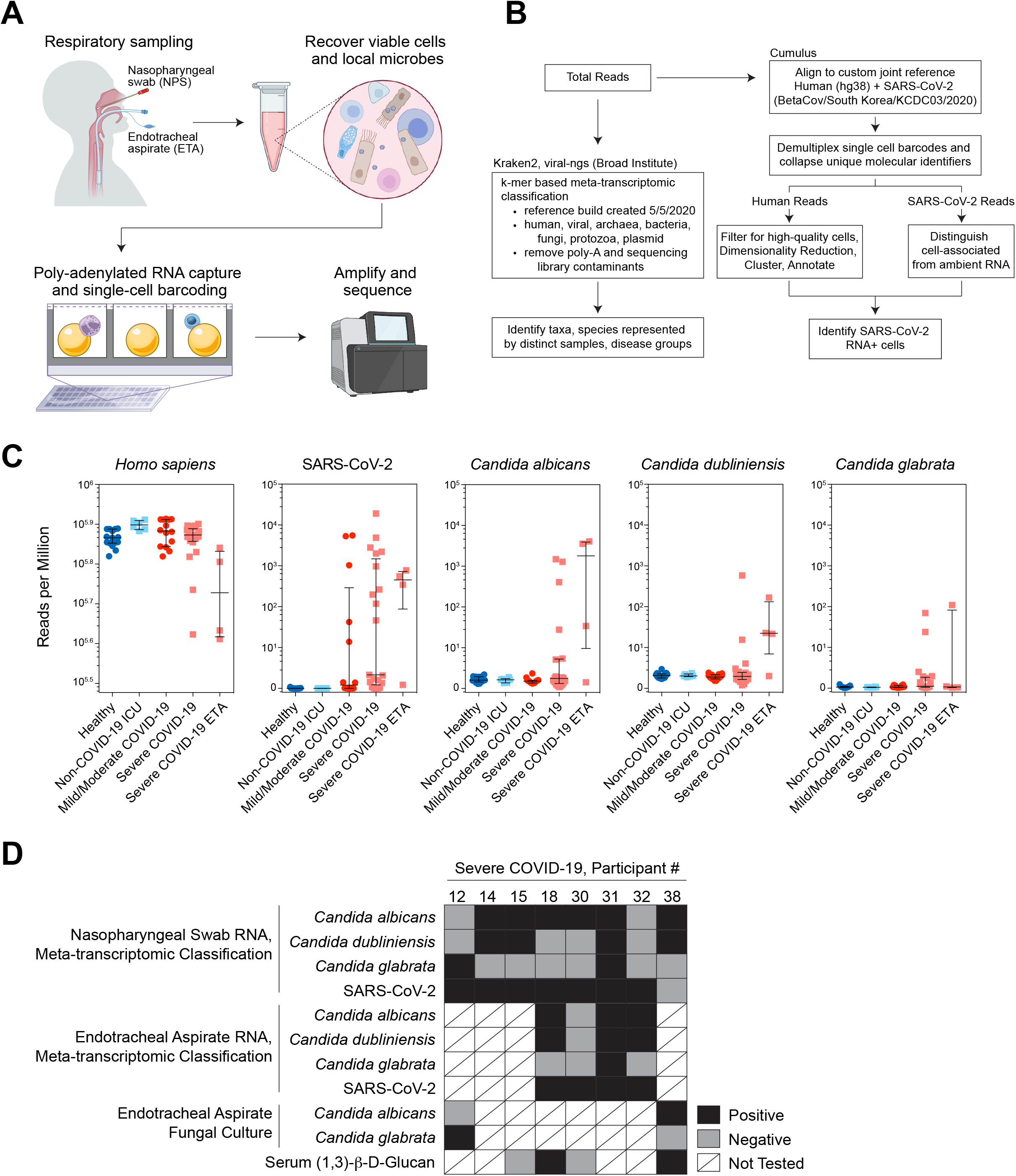
Co-detection of host single-cell transcriptome with intracellular and microenvironmental pathogen-derived genomic material. **A**. Schematic of biological sample processing pipeline **B**. Schematic of computational pipeline for each sample **C**. Abundance of human, SARS-CoV-2, and *Candida spp*. by participant and disease group as defined by meta-transcriptomic classification. N=56 participants. Lines represent median +/- interquartile range. **D**. Summary of results from sequencing, fungal culture, and serum (1,3)- β-D-glucan assays from 8 participants with detectable *Candida* species reads by nasopharyngeal swab or endotracheal aspirate.

Using meta-transcriptomic alignment of scRNA-seq data, we identified additional high-abundance microbial taxa across healthy participants and those with COVID-19 including common commensal microbes such as *Cutibacterium acnes, Malassezia restricta*, and *Staphylococcus aureus* (**Supplemental Figure 1A, Supplemental Table 1**). After SARS-CoV-2, the second-most abundant microbe detected was *Candida albicans*, which was detected on six NP samples obtained from patients with severe COVID-19 (**Figures 1C, 1D, Supplemental Figure 1A**). We also identified high levels of *Candida glabrata* in NP samples from 2 patients with severe COVID-19, and *Candida dubliniensis* in 4 samples. All samples that were positive for *C. glabrata* or *C. dubliniensis* were also positive for *C. albicans* with the exception of the NP sample obtained from COVID-19 participant 12. *Candida albicans* was also detected in 3 of 4 ETA samples obtained from patients with severe COVID-19. For one patient (COVID-19 participant 32) *C. albicans* was detected via ETA, but was not detected on their matched NP swab. All 3 ETA samples that were positive for *C. albicans* were also positive for *C. dubliniensis*, and one of these was also positive for *C. glabrata* (**Figures 1C, 1D**, and **Supplemental Figures 1A, 1B**). Notably, no *Candida spp*. or other fungal pathogens were detected within samples obtained from healthy individuals, those obtained from individuals with mild/moderate COVID-19, or those obtained from SARS-CoV-2 negative intubated patients in the intensive care unit with severe respiratory failure due to alternative causes. Thus, all *Candida spp*. reads were detected among patients who developed severe COVID-19 requiring intubation and mechanical ventilation (WHO severity score of 6-8).

Nearly all NP or ETA samples that were positive for *Candida spp*. were collected within one week of hospital admission (**Supplemental Figure 2A**). The majority (6/8) had been intubated for at least 1 day and 5/8 had received at least 1 day of corticosteroid treatment prior to sample collection. Clinical evaluations during hospitalization were performed to search for possible fungal infection for 3 of the 8 patients with high abundances of *Candida spp*. by meta-transcriptomic classification. COVID-19 participant 12, who had high abundance of *Candida glabrata* RNA sampled by NP swab on hospital day 2, was intubated on hospital day 1 and ETA fungal cultures sampled on hospital day 2 revealed growth of *Candida glabrata* (**Figure 1D**). For two participants, detection of *Candida spp*.-derived RNA via NP/ETA sampling significantly preceded clinical diagnostic testing for fungal pathogens. COVID-19 participant 38, whose NP swab revealed both *Candida albicans* and *Candida dubliniensis* RNA on hospital day 8, tested positive for serum (1,3)-β-D-glucan on hospital day 14 and had *Candida albicans* growth from ETA culture on hospital day 16. Both the NP and ETA samples from COVID-19 participant 18 obtained on hospital day 6 contained high abundances of reads classified as *C. albicans*, and *C. dubliniensis* was detected in the ETA sample. On hospital day 12, (1,3)-β-D-glucan was detected in this individual’s serum, prompting treatment with micafungin (an echinocandin antifungal).^23^ Together, this demonstrates that for a subset of patients, there was significant clinical concern during their hospitalization to prompt additional testing to evaluate for Candida respiratory infection or fungemia. Among those individuals, we find concordance between detection of *Candida* derived RNA from scRNA-seq libraries and clinical assays during their hospitalization.

We did not detect a difference in demographics or clinical characteristics – apart from severity of COVID-19 – between patients whose samples did or did not contain *Candida*-specific reads (**Table 1, Supplemental Figure 2B-2G**). 28-day mortality rates among individuals with severe COVID-19 were similar between *Candida spp*. positive vs. negative groups: 62.5% (5/8) among participants with *Candida spp*. detected, compared to 84.6% (11/13) among participants without *Candida* spp. detected. Nearly all (7/8) participants with COVID-19 whose samples contained *Candida spp*.-aligning RNA were previously diagnosed with type 2 diabetes mellitus (T2DM), and 8/8 were diagnosed with chronic hypertension. Notably, although T2DM represents an independent risk factor for mucosal *Candida* colonization, we did not find *Candida* or other fungal species among individuals with T2DM within the Healthy, non-COVID-19 intubated, or COVID-19 mild/moderate groups.^24^ Additionally, for individuals with recently measured HbA1c, we did not identify significant differences in the degree of glycemic control between individuals with different COVID-19 severity or by detection of *Candida*-specific reads (**Supplemental Figure 2G**).^25^ Critically, this analysis is based on a limited sample size, and merits further investigation with adequately-powered cohorts.

**Table 1.** Demographics and medical comorbidities of patients with severe COVID-19.

|  | COVID-19 severe<br>(WHO score 6-8)<br>*Candida spp.*<br>negative | COVID-19 severe<br>(WHO score 6-8)<br>*Candida spp.*<br>positive |
| --- | --- | --- |
| Case number | 23.2% (13/56) | 14.3% (8/56) |
| Age (years) |  |  |
| Minimum | 28 | 38 |
| Median (IQR) | 63 (49) | 57 (54.3) |
| Maximum | 79 | 84 |
| Sex |  |  |
| Female | 38.5% (5/13) | 62.5% (5/8) |
| Male | 61.5% 8/13 | 37.5% (3/8) |
| Ethnicity |  |  |
| Hispanic | 7.7% (1/13) | 0% (0/13) |
| Not Hispanic | 92.3% (12/13) | 100% (13/13) |
| Race |  |  |
| Black/African American | 53.8% (7/13) | 75% (6/8) |
| White | 23.1% (3/13) | 25% (2/8) |
| American Indian | 23.1% (3/13) | 0% (0/8) |
| BMI |  |  |
| Median (IQR) | 29.9 (27.8) | 37.0 (33.6) |
| Pre-existing conditions |  |  |
| Diabetes | 61.5% (8/13) | 87.5% (7/8) |
| Chronic kidney disease | 15.4% (2/13) | 25% (2/8) |
| Congestive heart failure | 7.7% (1/13) | 0% (0/8) |
| Lung disorder | 30.1% (4/13) | 50% (4/8) |
| Hypertension * | 69.2% (9/13) | 100% (8/8) |
| IBD | 0% (0/13) | 0% (0/8) |
| Treatment |  |  |
| Corticosteroids | 61.5% (8/13) | 75% (6/8) |
| Remdesivir | 7.7% (1/13) | 0% (0/8) |
| 28-day mortality *** | 84.6% (11/13) | 62.5% (5/8) |

IQR: inter-quartile range; BMI: body mass index; IBD: inflammatory bowel disease

Given the high frequency of *Candida spp*. colonization and clinically-relevant infection among individuals who developed severe COVID-19, we wondered whether the nasal mucosa of these individuals exhibits evidence for reactive or aberrant IL-17 responses. To better define the response of the human nasal epithelium to IL-17 we reanalyzed our previously-published population RNA-seq data which reflects gene expression in human nasal epithelial cells following *in vitro* exposure to a range of doses of IL17A (**Supplemental Figure 3**).^26,27^ Across multiple human donors, IL17A exposure led to upregulation of genes involved in keratinization (*SPRR2E, SPRR2F*, and *SPRR2G*), chemoattractant cytokines for lymphocytes, monocytes, and neutrophils (*CCL20, CXCL1, CXCL2*, and *CXCL3*), and pro-inflammatory factors such as *S100A7* and *S100A8* (**Figure 2A, Supplemental Figure 3A**).^28–30^ IL17A additionally resulted in dose-dependent induction of serum amyloid A genes *SAA1* and *SAA2* from nasal epithelial cells, which has previously been linked to pathogenic Th17 responses at barrier tissues.^31^

**Figure 2.**
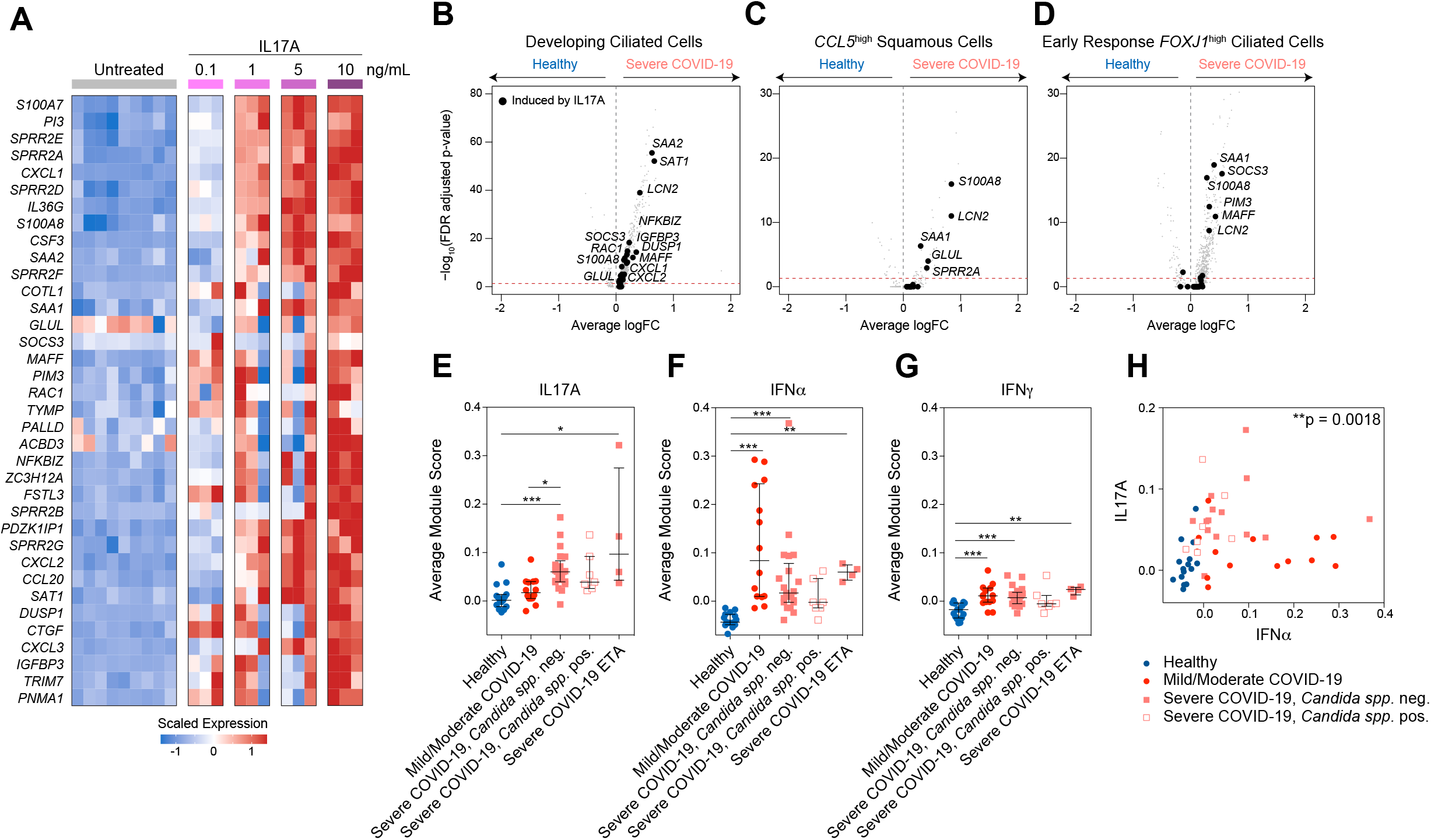
Respiratory epithelial transcriptional signatures following *in vitro* IL17A stimulation and *in vivo* fungal colonization in a severe COVID-19 cohort. **A**. Heatmap of population RNA-seq data comparing untreated nasal epithelial cells to those treated with increasing concentrations of IL17A as indicated across columns. Genes (rows) with significant differential expression between untreated and IL17A-treated conditions (FDR-corrected p < 0.05). **B.-D**. Volcano plots of differentially expressed genes between select epithelial cell types from healthy participants vs. those with severe COVID-19: developing ciliated cells (**B**), *CCL5*^high^ squamous cells (**C**), and early response *FOXJ1*^high^ ciliated cells (**D**). Grey points: all genes. Black points: genes induced in human nasal epithelial cells following IL17A treatment (as in **Figure 2A**). **E.-G**. Average gene module scores calculated for each participant, separated by disease group. Module score expression was computed over all epithelial cells. Input module genes derived from *in vitro* stimulation with each labeled cytokine: IL17A (**E**), IFNα (**F**), IFNγ (**G**). Statistical testing by Kruskal-Wallis test with Dunn’s post-hoc testing. *** p < 0.001, ** p < 0.01, * p < 0.05. Lines represent mean +/- SEM. **H**. Average gene module scores by participant, IFNα module on x-axis, IL17A module on y-axis. Points only reflecting NP samples. Point shapes and colors by disease group: Healthy: dark blue circles; Mild/Moderate COVID-19: red circles; Severe COVID-19, *Candida spp*. neg.: pink filled squares; Severe COVID-19, *Candida spp*. pos.: pink outlined squares. Statistical testing by Spearman’s correlation over all points: rho = 0.43, **p = 0.0018.

Using this RNA-seq data, we generated consensus gene sets for each tested cytokine that were robust across distinct donors, thereby giving us cell-type specific gene expression modules for IL17A, IFNα, IFNγ, IL1β, and IL4 (**Supplemental Figure 3, Supplemental Table 2**). Next, we returned to the scRNA-seq data from our human COVID-19 cohort and evaluated epithelial cells for transcriptomic signatures consistent with exposure to each cytokine. Compared to epithelial cells isolated from healthy controls, epithelial cells isolated from individuals with severe COVID-19 expressed significantly higher levels of genes that were also induced by IL17A exposure *in vitro* (**Figures 2B-D, Supplemental Figure 3B**). In particular, IL17A-induced genes *SAA1, SAA2, SAT1, LCN2, S100A8*, and *GLUL* were repeatedly significantly upregulated among diverse epithelial cell subsets in severe COVID-19, but were not found to be significantly induced within the nasal epithelia of patients with milder COVID-19. We confirmed that treatment of nasal epithelial cells with other inflammatory signals potentially found within the respiratory epithelium, including IL4 and IL1β, do not appreciably induce these factors, suggesting that induction of this gene module is a specific downstream effect of IL17 sensing (**Supplemental Figure 3C**).

Next, we directly scored epithelial cells for expression of gene signatures indicative of IL17A, IFNα, or IFNγ. (**Figures 2E-G**). Individuals who developed severe COVID-19 expressed consistently higher abundances of IL17A-induced genes compared to healthy participants or those with mild/moderate COVID-19 (**Figure 2E**). Interestingly, we did not detect higher abundances of genes in the IL17A gene module among participants with severe COVID-19 and detectable *Candida spp*. compared to participants with severe COVID-19 without detectable fungal pathogens. Likewise, we did not detect significant differences in levels of interferon-induced signatures between individuals with and without *Candida spp*. detected (**Figures 2F, 2G**). On an individual level, participants whose nasal epithelial cells expressed higher abundances of IFNα-induced genes did not express IL17A-induced genes, and vice versa (**Figure 2H**). Together, this suggests that induction of IL17A-stimulated genes in the nasal epithelium represents a shared feature of individuals who develop severe COVID-19, and is correlated with the absence of robust interferon-induced anti-viral responses.

## DISCUSSION

Our data demonstrate *Candida spp*. colonization of the upper respiratory tract in a significant proportion (38%) of individuals hospitalized with severe COVID-19 in a cohort from the University of Mississippi Medical Center sampled during the summer 2020 COVID-19 peak. Notably, no fungal pathogens were identified among individuals with mild or moderate COVID-19, non-COVID-19 ICU patients, or healthy controls. Fungal reads were detected by NP swab at early timepoints following hospital admission and within the acute phase of patients COVID-19 disease trajectory, suggesting that in some patients, fungal colonization and infection likely occurred prior to hospital admission and in advance of nosocomial exposures. *Candida* is not typically present in the nares of healthy people, being more readily detected in the oropharynx, thus identification of *Candida* from NP swabs of a subset of patients with severe COVID-19 would suggest either that severe COVID-19 predisposes to ectopic colonization in some hosts, or alternatively that our methodology is detecting increased fungal abundances derived from the oral mucosa^32^. Further direct comparisons of colonization in the mouth and nose of patients with severe COVID-19 will be necessary to clarify this issue.^33^

Our analysis additionally unites the use of single-cell transcriptomic technologies in human clinical cohorts with emerging computational approaches for meta-genomic pathogen classification of human samples, all derived from limited cellular material captured on a single NP swab^21,22^. By linking unbiased pathogen detection with single-cell nasal epithelial and immune transcriptional profiles, we have identified specific host behaviors indicative of response to a fungal pathogen and IL-17 signaling. For a subset of patients, early detection of *Candida spp*. in the upper respiratory mucosa corresponded to development of more extensive colonization and clinical concern for secondary fungal pneumonia and/or candidemia. Mounting evidence across various clinical cohorts, including our prior work, suggests that severe COVID-19 arises in individuals with impaired anti-viral immunity.^3,34–41^ While there is a paucity of evidence to suggest that type I/III interferon signaling directly restricts fungal colonization, prior *in vitro* studies indicate IL-17 signaling among airway epithelial cells may attenuate cellular responses to type I/III interferon^20,42–45^. Additionally, enhanced virally-induced epithelial injury resulting from impaired IFN signaling could facilitate *Candida* colonization.^46,47^ Surprisingly, we observe that IL17A-induced gene sets are elevated among epithelial cells from *all* individuals with severe COVID-19 in our cohort, even those patients without genomic or clinical evidence for co-incident fungal infection. Crucially, our data does not allow us to determine whether our observation of elevated IL-17 responses in patients with severe COVID-19 *without* overt evidence for fungal colonization is 1) the result of colonization below levels of detection in these assays, or 2) suggest that IL17 elevation represents a general phenotype of epithelial cells of patients with severe COVID-19, independent of fungal colonization. Future experiments encompassing longitudinal sampling of patients with COVID-19 could shed additional light on whether variability in the dynamics of IL-17 and interferon signaling may underlie *Candida* colonization in the upper airways.

Together, our data suggests upper respiratory Candida colonization and infection represents an underappreciated phenomenon among patients with severe COVID-19. Further research with larger cohorts is warranted to understand the frequency and timing of co-occurring infection with *Candida spp*. and other fungal pathogens following SARS-CoV-2 infection. Further, our data suggests that dedicated, multi-institutional studies are required to disentangle how clinical and subclinical fungal infections impact patient outcomes during hospitalization for COVID-19, and may hold key insights into determinants of severe respiratory failure for these patients and new strategies for diagnostic or therapeutic intervention.

## Data Availability

All data analyzed in this study is publicly available from prior manuscripts. Aligned and annotated scRNA-seq data can be downloaded via the Single Cell Portal, study SCP1289 (https://singlecell.broadinstitute.org/single_cell/study/SCP1289). Aligned TPM-normalized RNA-seq data can be downloaded via the Single Cell Portal, study SCP822 (https://singlecell.broadinstitute.org/single_cell/study/SCP822).

## ACKNOWLEDGEMENTS

We thank the study participants and their families for enabling this research, the clinical support staff at UMMC for assistance in sample collection, and the members of the Shalek, Ordovas-Montanes, Horwitz, and Glover labs for thoughtful discussion and comments on the project. This project has been made possible in part by grant number 2020-216949 from the Chan Zuckerberg Initiative DAF, an advised fund of Silicon Valley Community Foundation to A.K.S. and J.O.M.. C.G.K.Z. was supported by award number T32GM007753 and T32GM144273 from the National Institute of General Medical Sciences. J.O.M is a New York Stem Cell Foundation – Robertson Investigator. J.O.M was supported by the Richard and Susan Smith Family Foundation, the AGA Research Foundation’s AGA-Takeda Pharmaceuticals Research Scholar Award in IBD – AGA2020-13-01, the HDDC Pilot and Feasibility P30 DK034854, the Food Allergy Science Initiative, the Leona M. and Harry B. Helmsley Charitable Trust, The Pew Charitable Trusts Biomedical Scholars, The Broad Next Generation Award, The Chan Zuckerberg Initiative Pediatric Networks, The Mathers Foundation, The New York Stem Cell Foundation, and The Manton Foundation Cell Discovery Network at Boston Children’s Hospital. B.H.H. was supported by DK122532-01A1, NIH; 12019PG-CD002, The Leona M. and Harry B. Helmsley Charitable Trust; SRA #54518, and Crohn’s and Colitis Foundation. A.K.S. was supported by the Bill and Melinda Gates Foundation, Sloan Fellowship in Chemistry, the NIH (5U24AI118672), and the Ragon Institute of MGH, MIT and Harvard. This work was supported in part by the Division of Intramural Research of the NIAID.

## AUTHOR CONTRIBUTIONS

Conceptualization: J.O.-M., A.K.S., S.C.G., B.H.H.

Methodology: C.G.K.Z., A.H.O., S.C.G., J.O.-M., A.K.S., B.H.H.

Formal analysis: C.G.K.Z., A.H.O.

Investigation: C.G.K.Z., A.H.O.

Resources: S.C.G., A.H.O., M.Sl., H.L., A.P., C.B.S., M.W.B., M.Se., M.H., G.E.A.

Data Curation: T.O.R., Y.T., M.Sl. A.H.O., H.L., H.B.W., T.C., C.G.K.Z.

Writing – Original Draft: C.G.K.Z., A.H.O., J.O.-M., A.K.S., S.C.G., B.H.H.

Writing – Review & Editing: C.G.K.Z., V.N.M., A.H.O., A.W.N., Y.T., J.D.B., P.L., M.Sl., H.L., H.B.W., M.G., R.S.D., T.C., A.P., C.B.S., M.W.B., Y.P., M.H., G.E.A., M.Se., T.O.R., A.K.S., S.C.G., B.H.H., J.O.-M.

Visualization: C.G.K.Z.

Supervision: J.O.-M., A.K.S., S.C.G., B.H.H., T.O.R.

Project Administration: Y.P., T.O.R.

Funding Acquisition: J.O.-M., A.K.S., S.C.G., B.H.H.

## METHODS

### Participant Recruitment and Respiratory Sampling

Full participant characteristics are provided in previously published study^3^. The UMMC Institutional Review Board approved the study under IRB#2020-0065. All participants or their legally authorized representative provided written informed consent. Briefly, participants were eligible for inclusion in the COVID-19 group if they were at least 18 years old, had a positive nasopharyngeal swab for SARS-CoV-2 by PCR, had COVID-19 related symptoms including fever, chills, cough, shortness of breath, and sore throat, and weighed more than 110 lb. Participants were eligible for inclusion in the Healthy group if they were at least 18 years old, had a current negative SARS-CoV-2 test (PCR or rapid antigen test), and weighed more than 110 lb. COVID-19 participants were classified according to the 8-level ordinal scale proposed by the WHO representing their peak clinical severity and level of respiratory support required. Nasopharyngeal samples and endotracheal aspirate samples were collected by a trained healthcare provider, all processing and handling was carried out as previously described.^3,26^

### scRNA-seq Data Generation and Alignment

Annotated scRNA-seq data was recovered from Single Cell Portal (see **Data Availability** below) and single cell annotations were used as described in Ziegler et al. *Cell* 2021. Briefly, data represent aligned scRNA-seq libraries generated using Seq-Well S^3^, libraries were generated using Illumina Nextera XT Library Prep Kits and sequenced on NextSeq 500/550 High Output 75 cycle v2.5 kits to an average depth of 180 million aligned reads per array: read 1: 21 (cell barcode, UMI), read 2: 50 (digital gene expression), index 1: 8 (N700 barcode).^48^ Libraries were aligned using STAR within the Drop-Seq Computational Protocol (https://github.com/broadinstitute/Drop-seq) and implemented on Cumulus (https://cumulus.readthedocs.io/en/latest/drop_seq.html, snapshot 9, default parameters).^49,50^ As previously described, data were aligned using a custom reference which combined human GRCh38 (from CellRanger version 3.0.0, Ensembl 93) and SARS-CoV-2 RNA genomes.^3,51^

### Meta-Transcriptomic Pathogen Classification

To identify co-detected microbial taxa present in the cell-associated or ambient RNA of nasopharyngeal swabs, we used the Kraken2 software implemented using the Broad Institute viral-ngs pipelines on Terra (https://github.com/broadinstitute/viral-pipelines/tree/master).^21,22^ A previously-published reference database included human, archaea, bacteria, plasmid, viral, fungi, and protozoa species and was constructed on May 5, 2020, therefore included sequences belonging to the novel SARS-CoV-2 virus. Inputs to Kraken2 were: kraken2_db_tgz = “gs://pathogen-public-dbs/v1/kraken2-broad-20200505.tar.zst”, krona_taxonomy_db_kraken2_tgz = “gs://pathogen-public-dbs/v1/krona.taxonomy-20200505.tab.zst”, ncbi_taxdump_tgz = “gs://pathogen-public-dbs/v1/taxdump-20200505.tar.gz”, trim_clip_db = “gs://pathogen-public-dbs/v0/contaminants.clip_db.fasta” and spikein_db = “gs://pathogen-public-dbs/v0/ERCC_96_nopolyA.fasta”. Results were collected using the merge_metagenomics tool (https://viral-pipelines.readthedocs.io/en/latest/merge_metagenomics.html), and analysis and visualization of each samples’ metagenomic alignments was implemented in Prism (v6) or R (v4.0.2; packages ggplot2 (v3.3.2), Seurat (v3.2.2), ComplexHeatmap (v2.7.3)). All classification data is included as **Supplemental Table 1**.

### Human Nasal Epithelial Cell Response to *In Vitro* Cytokine Exposure

Gene lists representing human nasal epithelial cell responses to various exogenous cytokines *in vitro* are derived from a previously-published population RNA-seq data.^26,27^ Briefly, human nasal epithelial basal cells from 2 donors were stimulated *in vitro* with 0.1-10 ng/mL IFNα, IL17A, IFNγ, IL1β, or IL4 for 12 hours. Following stimulation, cells were lysed and bulk RNA-seq libraries were generated using the SMART-Seq2 protocol.^52^ We identified epithelial gene sets induced by each cytokine independently by testing for differentially expressed genes compared to matched, untreated nasal epithelial samples (n=10). Differential expression testing was carried out using a likelihood ratio test assuming a negative binomial distribution, implemented with the Seurat (v3.1.5) FindAllMarkers function (test.use = “negbinom”). We considered genes as differentially expressed with an FDR-adjusted p value < 0.05. ^53^

To score for cytokine-specific gene expression among COVID-19 or Healthy scRNA-seq samples, we first subsetted our scRNA-seq data to only epithelial cells using “Coarse” cell types, as defined by cell typing procedure carried out in prior publication (Ziegler et al. *Cell* 2021). Coarse cell type groups that were included in the analysis: “Ciliated Cells”, “Developing Ciliated Cells”, “Secretory Cells”, “Goblet Cells”, “Ionocytes”, “Deuterosomal Cells”, “Squamous Cells”, “Basal Cells”, “Mitotic Basal Cells”, and “Developing Secretory and Goblet Cells”. We calculated module scores over all epithelial cells using the Seurat function AddModuleScore with default inputs. The average module score for each NP or ETA sample was utilized as a representative measure of epithelial behavior for each participant, as represented in **Figures 2E-2H**.

### scRNA-seq Analysis of Differential Expression

To compare gene expression between cells from distinct disease groups (e.g. Healthy vs. Severe COVID-19, *Candida spp*.), we employed a likelihood ratio test assuming a negative binomial distribution as described above (using Seurat FindAllMarkers function (test.use = “negbinom”)).^53^ We considered genes as differentially expressed with an FDR-adjusted p value < 0.001 and log fold change > 0.25. Results from select “Detailed” cell types, as defined and previously reported by Ziegler et al. are represented in **Figures 2B-D**.^3^ Full results of differential expression as represented in **Figures 2B-D** can be found in Supplementary Tables accompanying the published dataset.

### Statistical Testing

All statistical tests were implemented in R (v4.0.2) or Prism (v6) software. Specific statistical tests, p-values, n, and all summary statistics are provided in the results section, figure legends, and/or figure panels.

## Data Availability

All scRNA-seq and RNA-seq data analyzed in this study is publicly available from prior manuscripts. Aligned and annotated scRNA-seq data can be downloaded via the Single Cell Portal, study SCP1289 (https://singlecell.broadinstitute.org/single_cell/study/SCP1289). Aligned TPM-normalized RNA-seq data can be downloaded via the Single Cell Portal, study SCP822 (https://singlecell.broadinstitute.org/single_cell/study/SCP822). Results from Kraken2 meta-genomic classification are reported in **Supplemental Table 1**.

## SUPPLEMENTAL FIGURE TITLES AND LEGENDS

**Supplemental Figure 1. Meta-transcriptomic classification of reads from nasopharyngeal swabs and endotracheal aspirates**

**A**. Heatmap of top detected microbes (rows) across all samples (columns). Bar plot on left: total classified reads per million (M) for each microbe listed along heatmap rows. Heatmap colors represent total classified reads per M, dark red: higher abundance, yellow: lower abundance, white: < 10 reads per M.

**B**. SARS-CoV-2 and *Candida spp*. reads detected from matched nasopharyngeal (NP, black bars) and endotracheal aspirate (ETA, grey bars) samples from the same participants.

**Supplemental Figure 2. Participant characteristics and clinical course**

**A**. Hospital timelines for 8 participants with *Candida spp*. detected from scRNA-seq NP or ETA samples.

**B.-D**. Select categorical demographic and clinical information from study participants by disease cohort: 28-day mortality (**B**), type 2 diabetes mellitus (T2DM) (**C**), and chronic hypertension (**D**). Statistical testing by multi-group chi-square test, significance result is reported above each plot.

**E.-F**. Select continuous demographic and clinical information among participants with severe COVID-19, separated by detection of *Candida spp*.: age (**E**) and BMI (**F**). Statistical testing using student’s t-test. Lines represent median +/-interquartile range.

**G**. Hemoglobin A1c (HbA1c) for each study participant by disease cohort. Dashed line at HbA1c 6.5%. Group differences non-significant by Kruskal-Wallis test. Numbers in parenthesis reflect patients per group with available data in medical record.

**Supplemental Figure 3. Transcriptional responses of respiratory epithelial cells following *in vitro* exposure to various cytokines**

**A**. Expression of select genes following 12-hour stimulation with increasing doses of IL17A. Each gene significantly upregulated following IL17A expression by likelihood ratio test, FDR-adjusted p-value < 0.001. Lines represent mean +/-SEM.

**B**. Heatmap of IL17A-induced genes among developing ciliated cells from NP swabs. Heatmap colors reflect scaled gene expression: red: higher expression, blue: lower expression. Left columns (blue bar): developing ciliated cells from n=15 Healthy participants; right (pink bar): developing ciliated cells from n=8 participants with severe COVID-19 and *Candida spp*. detected. Genes (rows) selected represent genes significantly upregulated among cultured human nasal epithelia cells following *in vitro* exposure to IL17A. Statistical significance comparing Healthy-derived vs. Severe COVID-19, *Candida spp*. positive-derived single cells by likelihood ratio test assuming an underlying negative binomial distribution. *** FDR-corrected p < 0.001, ** FDR < 0.01, * FDR < 0.05.

**C**. Expression of select genes among human nasal basal cells by stimulation condition.

## SUPPLEMENTAL TABLES TITLES

**Supplemental Table 1. Raw meta-transcriptomic classification data from Kraken2**

**Supplemental Table 2. Gene lists for scoring epithelial responses to cytokines**

