## Supplementary figures and images for "Severe COVID-19 is associated with fungal colonization of the nasopharynx and potent induction of IL-17 responses in the nasal epithelium"

### Supplemental Figure 1

##### Supplemental Figure 1

A

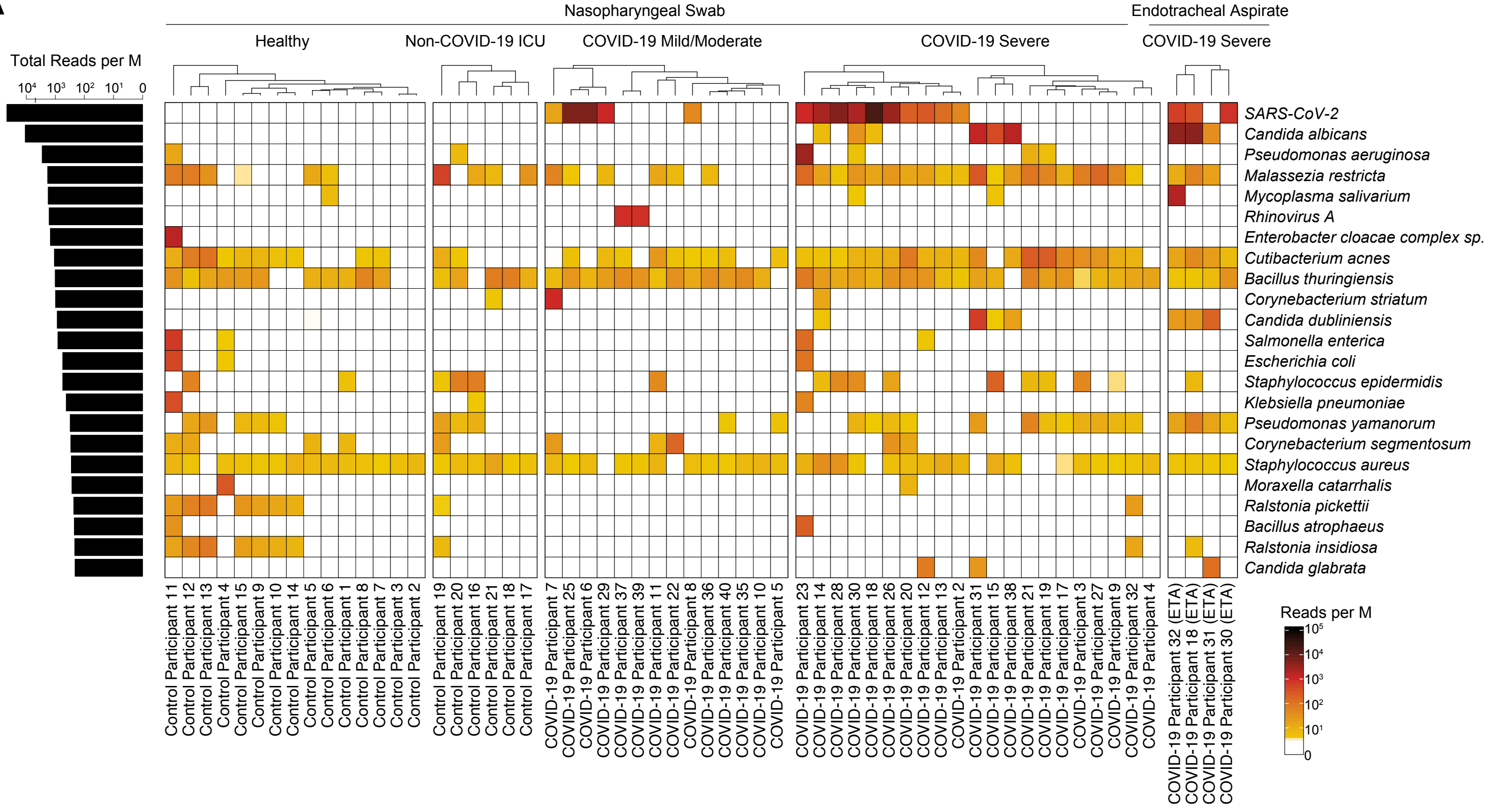


B

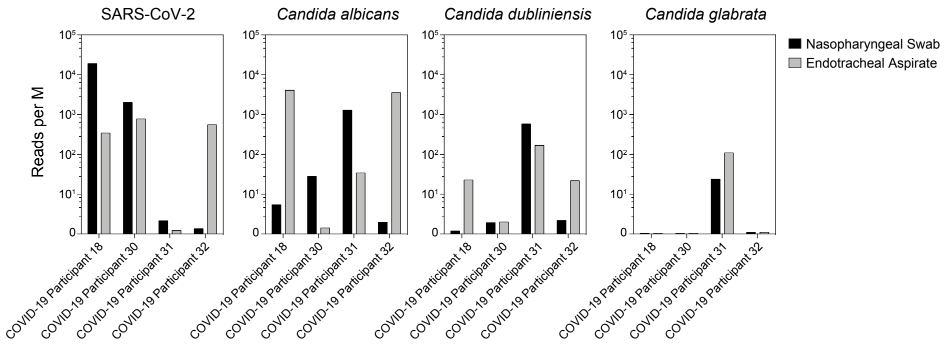

### Supplemental Figure 2

#### Supplemental Figure 2

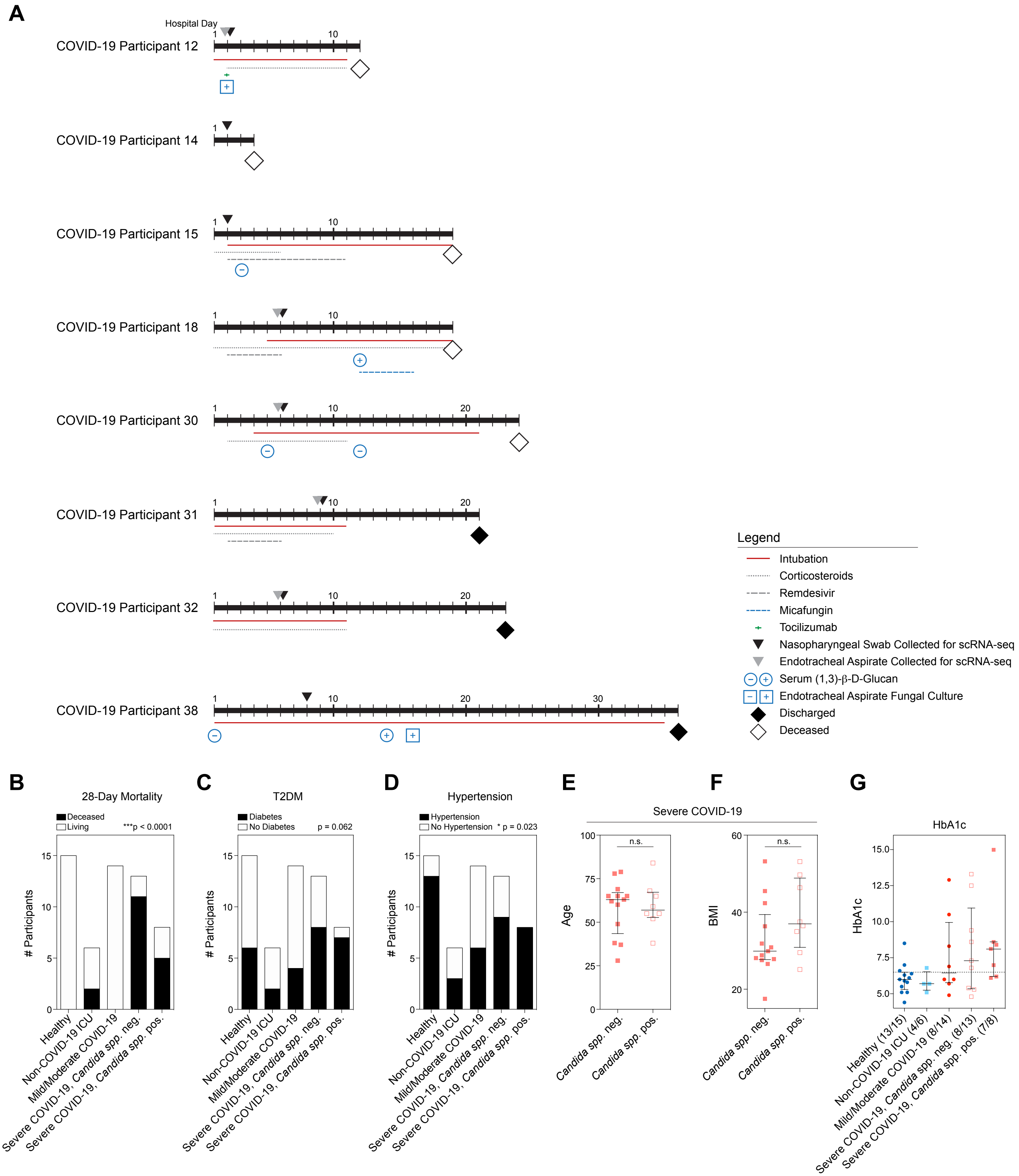

### Supplemental Figure 3

#### Supplemental Figure 3

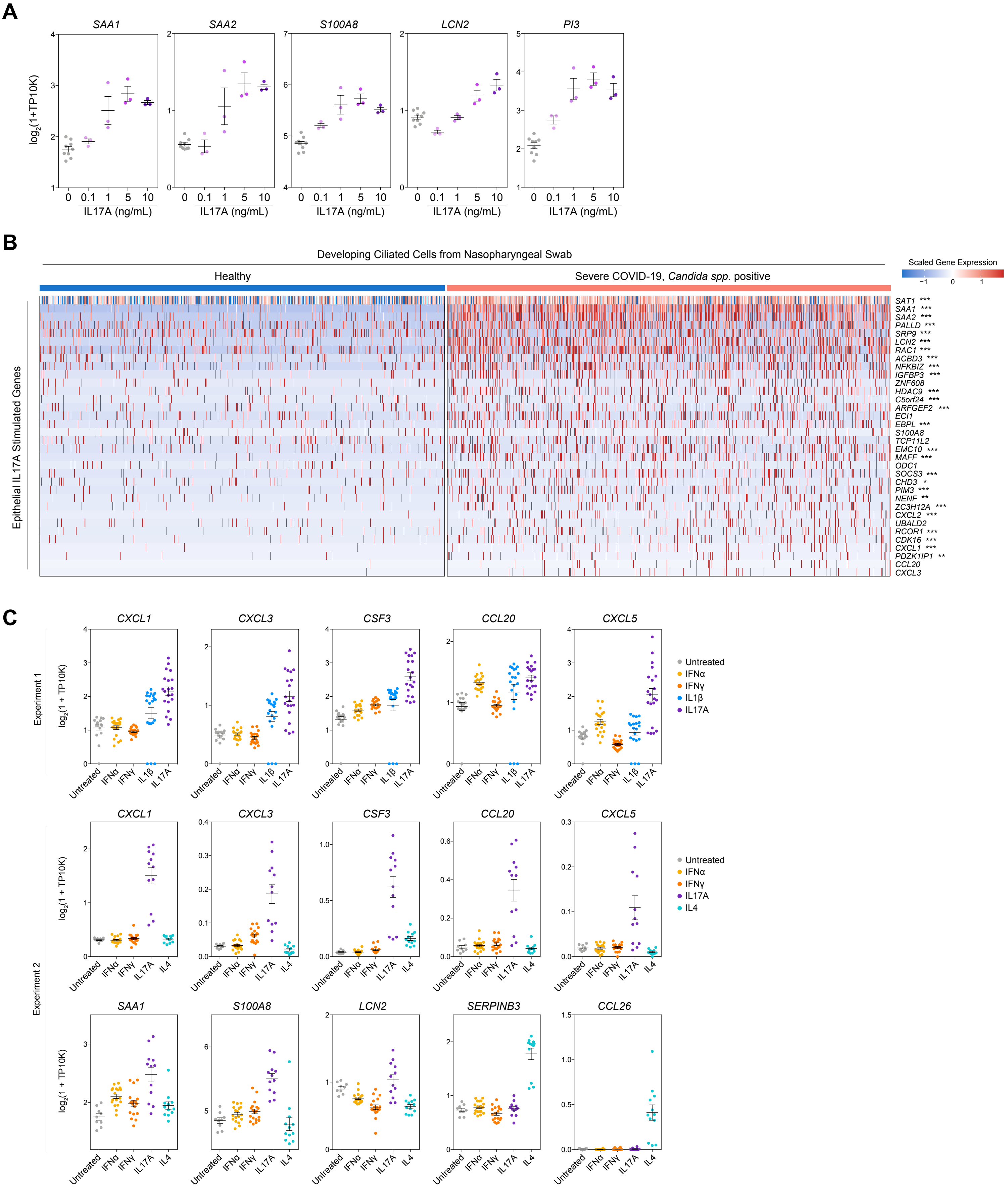
